# Risk Perception, Behavior, and Knowledge of Typhoid Fever Diagnosis, Treatment, and Vaccination: Insights from Patients in Kano, Nigeria

**DOI:** 10.1101/2024.11.04.24316691

**Authors:** Nirmal Ravi, Oluwaseunayo Deborah Ayando

## Abstract

**Background:** Typhoid fever, caused by *Salmonella enterica* serovar Typhi and *S. Paratyphi*, remains a major public health issue in Nigeria, exacerbated by limited access to clean water, inadequate sanitation, and a lack of awareness about vaccination. To inform future interventions, this study investigates patients’ knowledge and perceptions of typhoid fever, including its diagnosis, treatment, and prevention.

**Method:** A prospective cross-sectional study was conducted among adults attending EHA Clinics in Kano, Nigeria. A structured survey assessed participants’ knowledge of typhoid fever, its transmission, its symptoms, and their attitudes toward the typhoid vaccine. Data were analyzed using R software, with descriptive statistics and associations between variables examined.

**Results:** Out of 443 participants, 71.4% were aware of typhoid fever, with 98% expressing confidence in its curability through appropriate treatment. Awareness of the typhoid vaccine, however, was low at 31.1%. Risk perception was high, with 75.5% of participants likely to trust a positive test result, and 63.4% believed they had a chance of contracting typhoid within the next year. Although antibiotics were widely regarded as beneficial (51.9%), misconceptions about vaccine harm persisted among some of the participants. Socio-demographic factors, particularly gender and education, were significantly associated with vaccine interest, with males and those with lower education levels showing greater interest in vaccination.

**Conclusion:** Despite high awareness of typhoid fever, misconceptions and limited knowledge about the vaccine pose challenges for prevention in Nigeria. Targeted educational campaigns could address these gaps and promote better attitudes toward vaccination and preventive behaviors.

## Introduction

Enteric fever (typhoid and paratyphoid fever) is caused by Salmonella enterica serovar Typhi (S. Typhi) and Salmonella enterica serovar Paratyphi (S. Paratyphi). S. Paratyphi A and B (uncommonly, S. Paratyphi C) cause a disease clinically indistinguishable from typhoid fever, particularly in parts of Asia ^[1].^ Approximately 21.7 million new cases ^[2]^ and 21,7000 deaths occur as a result of the infection of typhoid fever yearly. Antillon et al. (2017) reported that approximately 17.8 million cases of the disease occur yearly in low-middle-income countries ^[2].^ Contaminated water is one of the disease’s transmission pathways ^[3].^ Human infection with Salmonella is mainly by the oral route through ingestion of fecal contaminated food and water, meat from infected animals, unclean hands, and house flies ^[4],^ Other risk factors include patronizing food vendors and a history of contact with a case or a chronic carrier, amongst others. Environmental factors such as the rainy season, open sewers, contaminated water bodies, and low-elevation areas have been implicated ^[5].^

Symptoms of the disease include prolonged high fever ^[6]^, fatigue, headache, nausea, abdominal pain, and constipation or diarrhea. Some patients may have a rash ^[6]^. Severe cases may lead to serious complications or even death ^[6]^. Typhoid fever can be treated with antibiotics however, resistance to common antimicrobials has become widespread, especially in endemic areas ^[6]^. An estimated 2.1– 6.5 million cases of Invasive non-typhoidal salmonella (iNTS) disease occur annually, with the highest incidence in Africa ^[7]^. The risk of iNTS is highest in infants, young children, and young adults with underlying comorbidities, including severe anemia, malaria, malnutrition, and HIV infection ^[1]^.

Typhoid vaccines have been available for disease control and prevention since 1896; however, their use as a routine tool for disease prevention in endemic settings has been hampered because of insufficient data on disease burden ^[8]^. Even though typhoid fever can be treated with antibiotics, treatment as an alternative to prevention has proven ineffective ^[9].^

Typhoid fever is a life-threatening disease ^[10]^ that has posed a public health concern in many parts of the world, especially low and middle-income countries for decades ^[6]^. In 2017, a global incidence of 14.3 million was recorded ^[11]^. In the same year, a global case fatality rate of 0.95% was estimated, with higher approximates among people living in lower-income countries ^[11]^ such as Nigeria. In Nigeria, typhoid fever incidence ranges from 3.9% to 18.6% ^[5]^, with the highest cases seen among young and middle-aged adults ^[4]^.

The typhoid vaccine has been proven to be an effective way of preventing the disease ^[9]^. However, its use as a standard tool for Disease prevention has been foiled, especially in endemic areas ^[9]^.

In addition to poor sanitation, lack of access to clean water, and poor toilet facilities; it has been found that misconception about typhoid is a risk factor for the disease ^[12]^ and contributes to its endemicity in affected countries such as Nigeria.

Though there are many studies on the prevalence of typhoid fever; no research has delved into the knowledge and perception of people towards typhoid vaccine in Nigeria.

This study, therefore, aims to add more information to the existing data on people’s knowledge of typhoid fever; and provide evidence on knowledge and perception of the typhoid vaccine among Nigerians to serve as a template for other researchers in the future. In the same vein, we hope to, based on our study findings, be able to make targeted recommendations aimed at promoting better attitudes towards the disease and its prevention including vaccination.

## Methods

**Study Area:** EHA Clinics is located in Kano, the municipal center of Kano state in Northcentral Nigeria. The municipality’s total area is 193 square miles and it has the second-largest population in Nigeria following Lagos. The urban population in 2018 was 3.8 million with an estimated growth rate of 3.1% from 2018 to 2030 [21]. Malaria transmission is meso-endemic in Kano State, Nigeria with a prevalence of 32% [22].

**Study setting and population:** This study was a prospective cross-sectional study conducted to give detailed information on the knowledge and attitudes of patients in EHA Clinics, Kano towards typhoid, typhoid diagnosis, treatment, and vaccine. The main target population was adults aged 18 and older attending EHA Clinics Kano at the time of the study.

**Eligibility criteria:** All adult patients aged 18 and above who consent to participate in the study. **Data collection:** A trained Research Associate was assigned to administer a structured survey, which targeted adults aged 18 and above visiting EHA Clinics in Kano. Participation in the study was voluntary, with all participants providing informed consent before the commencement of the survey.

**Scoring of Knowledge Questions:** Seven questions each were used to assess the knowledge of study participants on the mode of transmission and symptoms of typhoid fever. Study participants who gave the right response were given a score of 1 and those who did not were given a score of 0. Participants who got 5 or more questions correctly were considered to have good knowledge while participants with a score of 2 or less were considered to have poor knowledge

**Statistical analysis:** The data collected was analyzed using R software. Descriptive statistics were presented as tables, perception data was presented using charts and the association between categorical variables was explored using Fisher’s exact test. Kruskal-Wallis H Test was used to compare the perceived severity of typhoid among respondents who have had typhoid before vs those who have never had typhoid before and also to compare the perceived severity of typhoid among respondents whose family members have been admitted for typhoid treatment before vs those whose family members have never been admitted for typhoid before.

**Ethical considerations:** Ethical approval for the study was granted by the Kano State Ministry of Health Research Ethics Committee (approval number: NHREC/17/03//2018). All data were de-identified before analysis and the authors had all necessary administrative permissions to access the data.

**Informed consent:** Informed verbal consent was obtained from each respondent before the start of the questionnaire administration. The objectives, benefits, and risks of the study were explained in detail to each participant; and they were informed that the refusal to participate in the research or withdrawal, have no consequences.

## Results

### Socio-demographic of participants

A total of 443 participants were recruited for the study with 188 (42.4%) male and 255(57.6%) female. Most of the participants (86.3%) attended college and above. 47.8% of them were Hausa’s by tribe and 11.3% belonged to other tribes aside from the major ethnic groups in Nigeria. More than half of the participants were employed (59.4%) while 10.4% were unemployed.

**Table 1:** Socio-demographic of participants.

| **Variables** | **Frequency (%)** |
| --- | --- |
| **Gender (n=443)** |  |
| Male | 188 (42.4%) |
| Female | 255 (57.6%) |
| **Age distribution** | Mean age- 32.5 years |
| **Educational Qualification (n=438)** |  |
| College and above | 378 (86.3%) |
| Secondary school | 56 (12.8%) |
| Primary school | 3 (0.7%) |
| Never attended school | 1 (0.2%) |
| **Ethnicity (n=443)** |  |
| Hausa | 212 (47.8%) |
| Igbo | 27 (6.1%) |
| Yoruba | 50 (34.8%) |
| Other | 154 (11.3%) |
| **Employment status (n=443)** |  |
| Employed | 263 (59.4%) |
| Unemployed | 46 (10.4%) |
| Student | 68 (15.3%) |
| Retired | 10 (2.3%) |
| Other | 56 (12.6%) |
| **Perception of healthiness (n=441)** |  |
| Excellent | 87 (19.7%) |
| Very good | 178 (40.4%) |
| Good | 144 (32.7%) |
| Fair | 31 (7.0%) |
| Poor | 1 (0.2%) |

### Participants knowledge assessment

More than half of the participants have heard of typhoid fever (71.4%), and many believe it can be cured with appropriate treatment (98.0%).

Less than half of the participants were aware of the typhoid vaccine (31.1%) while 43.4% did not know about it. Generally, most participants have a good knowledge of typhoid transmission and symptoms; 97.5% and 98.6% respectively. A majority of the participants, 95.5% and 85.2% respectively, believed that drinking unclean water and not washing fruits/vegetables before eating them were significant risk factors for contracting typhoid fever.

**Table 2:** Knowledge Assessment of Participants.

| Variables | Frequency (%) |  |  |
| --- | --- | --- | --- |
|  | Yes | No | Not sure |
| Informed about typhoid fever | 317 (71.4%) | 127 (28.6%) | - |
| Typhoid can be cured with appropriate treatment | 434 (98.0%) | 3 (0.6%) | 6 (1.4%) |
| Aware of the typhoid vaccine | 137 (31.1%) | 192 (43.4%) | 112 (25.4%) |

### Participants’ perception of Typhoid fever severity compared with other diseases

The perception of typhoid severity compared with other diseases was assessed among participants as shown in Figure 1. The first most perceived to be a severe disease by the participants (76.6%) was breast cancer while the second most perceived to be a severe disease by the participants was diabetes (67.8%). Typhoid fever was considered very serious by 60.0% of the participants, while nearly half of them, 49.3%, perceived COVID-19 as a severe illness.

**Figure 1:**
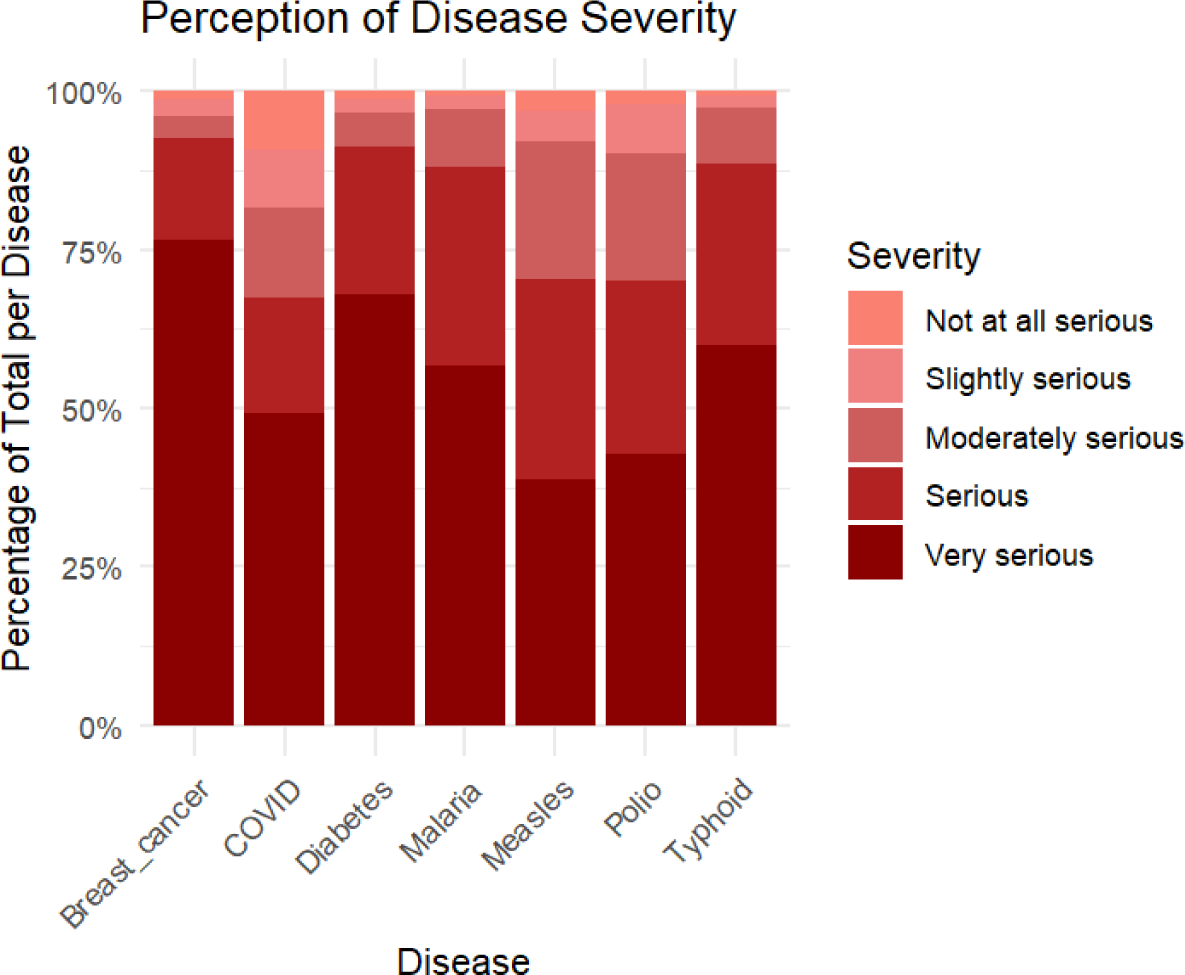
Participants’ perception of Typhoid fever severity compared with other diseases.

### Participants’ perception of the benefits of antibiotics, smoking, tea/coffee, and vaccines

As shown in Figure 2, the majority of the participants considered antibiotics to be beneficial (51.9%), 50.2% of them considered vaccines to be beneficial while most of them (91.7%) did not perceive smoking to have benefits to their health.

**Figure 2:**
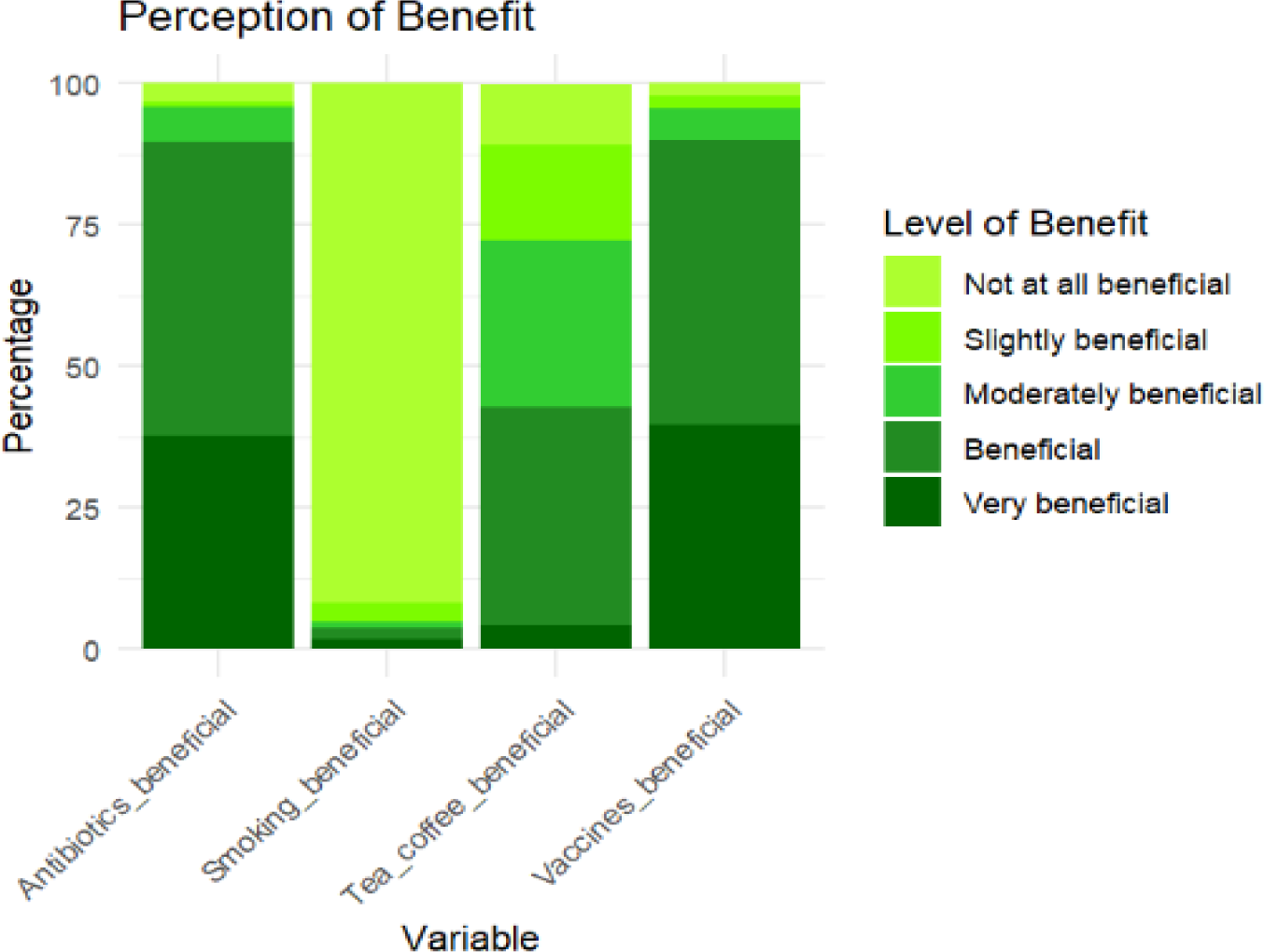
Participants’ perception of the benefits of antibiotics, smoking, tea/coffee, and vaccines.

### Participants’ perception of the harm of antibiotics, smoking, tea/coffee, and vaccines

This study shows that less than 5% of the participants believe that taking vaccines is harmful while the majority (67.3%) did not think it is harmful. Most of the participants (88.2%) perceived that smoking is very harmful to their health and a majority of them think taking tea/coffee is not at all harmful (52.1%).

**Figure 3:**
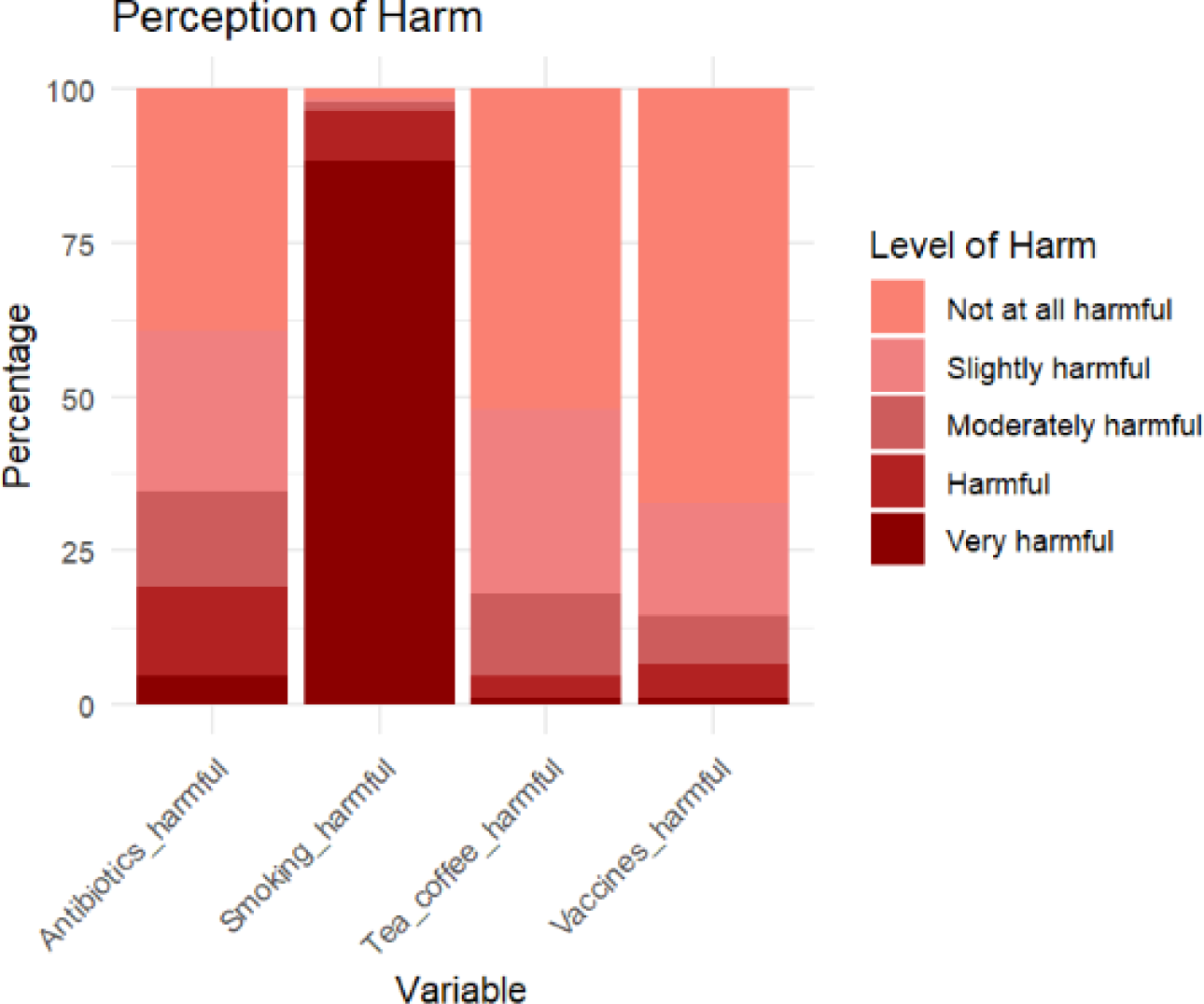
Participants’ perception of the harm of antibiotics, smoking, tea/coffee, and vaccines.

### Participants’ perception of diagnosis of typhoid fever and future risk

To assess the participants’ perception of typhoid fever diagnosis and risk of having typhoid, they were asked about the likelihood of believing a positive test result and their chance of having typhoid in the next year. Most participants, 75.5%, would believe their positive typhoid test results, while around 15% thought there was only a slight likelihood of this occurring. Additionally, fewer than 10% believed they could not test positive for typhoid when tested. More than half of the participants 63.4% believed there’s a likelihood of them having typhoid in the next year although about 22.3% of them think they only have a slight chance of having the disease.

**Figure 4:**
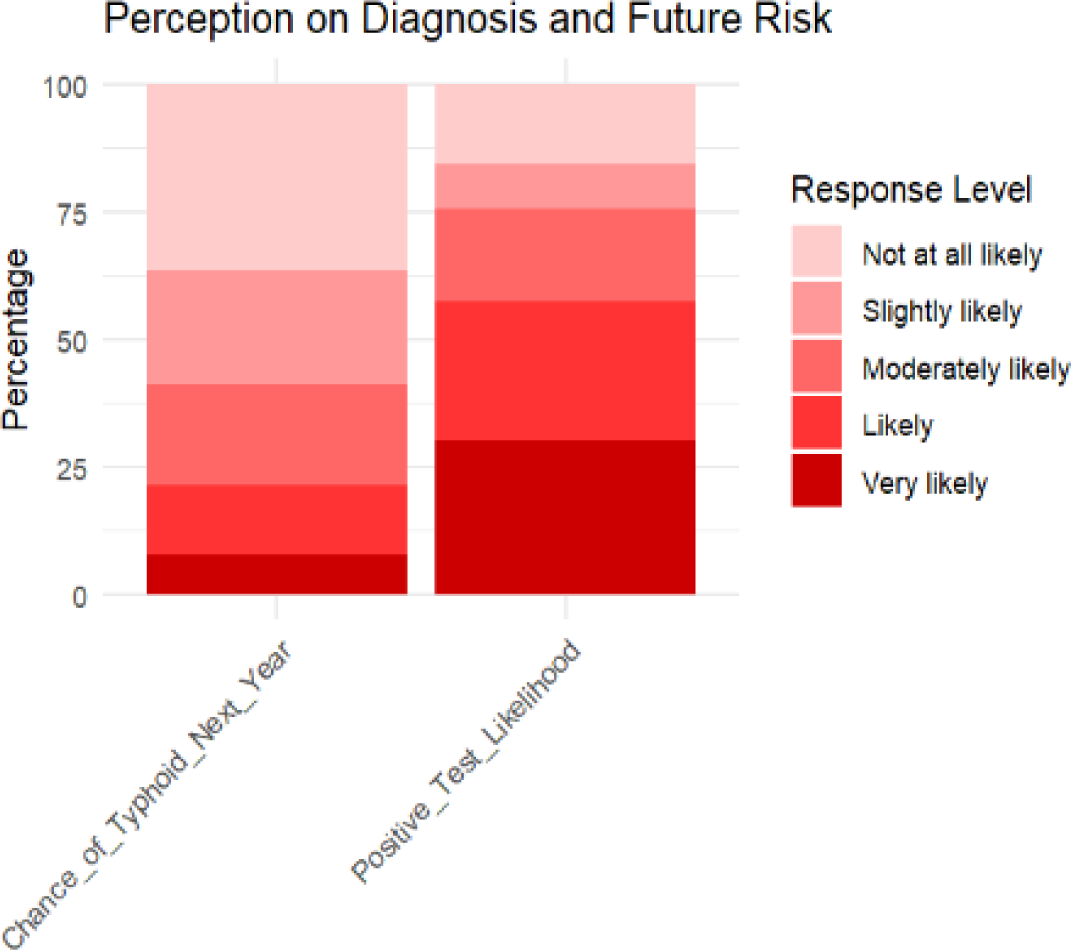
Participants’ perception of diagnosis of typhoid fever and future risk.

### Association between Socio-demographic Factors and Vaccine Uptake

The male participants have about 0.7% higher risk of being interested in the vaccine when compared to females when other factors are controlled. A risk ratio of 1.383 suggests that individuals who never attended school have a 38.3% higher risk of being interested in the vaccine when compared to the reference education category while a risk ratio of 0.828 among participants who attained secondary education indicates a 17.2% lower risk of being interested in the vaccine compared to the participants who attained higher education. Participants who were Students and Unemployed showed a lower of being interested in taking the vaccine with a risk ratio of 0.977 and 0.964 respectively while the retired individuals had a 4.3% higher risk of being interested when compared to the participants that are employed. A risk ratio of 0.856 among participants who were Igbos indicates a 14.4% lower risk of being interested in taking the vaccine while a risk ratio of 1.088 among the Yoruba participants indicates a slight increase in their interest in taking the vaccine when compared to the Hausa participants.

**Table 3:** Association between Socio-demographic Factors and Vaccine Uptake.

| Variables |  | RR | CI |
| --- | --- | --- | --- |
| Sex | Male | 1.0072552 | 0.183-0.204 |
| Level of education | Never attended school | 1.3834593 | -0.421 - 0.117 |
|  | Primary school | 1.0643612 | -0.388 - 0.407 |
|  | Secondary school | 0.8281820 | -0.346 - 0.152 |
| Employment status | Others | 0.8901559 | -0.158 - 0.326 |
|  | Retired | 1.0434134 | -0.474 - 0.089 |
|  | **Student** | 0.9765396 | -0.222 - 0.553 |
|  | **Unemployed** | 0.9618966 | -0.188 - 0.062 |
| **Ethnicity** | **Igbo** | 0.8562382 | -0.459 - 0.385 |
|  | **Yoruba** | 1.0868198 | -0.348 - 0.277 |
|  | **Others** | 1.0880187 | -0.161 - 0.099 |
| **Age** |  | 1.0033117 | 0.074 - 0.012 |

## Discussion

Our study found that 71.4% of participants were informed about typhoid fever. This finding is consistent with the findings from a qualitative study conducted by Ilouno George (2020), who reported high awareness levels of typhoid fever among participants in Nimo Village, Nigeria. Similarly, Mulu et al. (2021) found a comparable level of awareness (98.7%) among patients in Northwest Ethiopia, with a significant proportion recognizing the symptoms and transmission modes of typhoid fever. These consistent findings across different regions indicate a generally high level of awareness about typhoid fever in endemic areas, likely due to its widespread prevalence and the efforts of public health education campaigns.

The findings of our study revealed a relatively low awareness of the typhoid vaccine (31.1%), which is similar to the 22% awareness observed in India (Chetana, 2020) but contrasting with findings from two similar studies by Jamal et al. 2020 and Tahir et al. 2023 in Pakistan where 54.2% and 75% awareness were reported respectively. This may be attributed to the possibility that these studies were conducted in the vicinity of the vaccination center having the probability of raising consciousness and awareness about the vaccines. In addition, Blum et al. (2014) reported higher vaccine awareness in Malawi during a typhoid outbreak, attributing this to intensified public health campaigns. A study by Stanaway et al. (2019) indicated that vaccine awareness and uptake were significantly higher in South Asia, where targeted vaccination programs were implemented. Notably, a vast majority of participants (98%) believe that typhoid can be effectively treated with appropriate treatment, reflecting a high confidence level in medical interventions for the disease. This study also indicated that only half of the participants (47.8%) believed unclean water was a significant risk factor for typhoid fever. This is lower than the findings from a study among patients in Northwest Ethiopia, where 86.1% of the participants believed drinking contaminated water is a means of transmission of typhoid fever.

Regarding treatment perceptions, 98% of our study participants believed that typhoid fever could be cured with appropriate treatment, which aligns with global perspectives. According to the World Health Organization (2019), appropriate antibiotic treatment is effective for typhoid fever, although resistance to common antimicrobials is a growing concern. This high level of confidence in treatment underscores the importance of continuous monitoring of antibiotic resistance patterns and updating treatment guidelines accordingly.

Our study’s findings on the perceived severity of typhoid fever compared to other diseases revealed that 60% of participants considered typhoid fever very serious. This is in line with Pitzer et al. (2019), who discussed the hidden burden of typhoid fever compared to more visibly impactful diseases like cancer. However, a study by Enabulele and Awunor (2016) in Nigeria found that many healthcare workers perceived typhoid fever as less severe than other common infectious diseases, indicating variability in perceived severity depending on the population.

A study conducted by Guo et al. on risk perception, health efficacy, and health behaviour for cardiovascular disease applied the Risk Perception Attitude framework (RPA) that to categorize individuals based on their risk perception and efficacy beliefs. The RPA framework proposes that risk perception and efficacy belief not only have a direct impact but also a moderating effect on individuals’ health-related behavior (Rimal, 2003). It suggests that people can be categorized into one of four distinct groups based on risk perception and efficacy level. The “Responsive” category refers to individuals who have high levels of both perceived risk and self-efficacy; individuals with a high level of perceived risk but low self-efficacy were labeled as “Avoidant”; individuals with a low level of perceived risk but high self-efficacy were categorized into the “Proactive” group; and individuals with both a low level of perceived risk and self-efficacy were labeled as “Indifferent”. Higher risk perception was associated with increased engagement in preventive behaviors, provided individuals also have high efficacy beliefs (Guo et al., 2023). Therefore, improving risk perception through education about the severity and transmission of typhoid disease can encourage preventive behaviors such as vaccination and proper hygiene. Another study in the United Arab Emirates assessed Adolescents’ use of online food delivery applications and perceptions of healthy food options and food safety (Saleh et al., 2024). The study highlights the importance of efficacy beliefs in motivating individuals to make healthier food choices. Similar strategies can be applied to typhoid prevention by enhancing individuals’ confidence in their ability to prevent infection through proper practices. In addition, a qualitative study explored perceptions, attitudes, and practices of adolescent university students in Lagos, Nigeria, towards a healthy lifestyle. The study revealed that most participants understood a healthy lifestyle to involve a balanced diet, regular physical activity, and disease prevention (Menakaya & Menakaya, 2022). This understanding can be linked to our study, where knowledge and awareness of transmission pathways and risk factors are critical for disease prevention. Participants’ attitudes towards a healthy lifestyle were shaped by personal conviction and disposition. Individuals’ attitudes towards typhoid disease, including misconceptions and lack of awareness, could influence their preventive behaviors, however, this was not assessed in our study.

The association between socio-demographic factors and vaccine uptake in our study indicated that males, individuals with lower education levels, and retired participants were more interested in the vaccine. This trend contradicts findings from Khan et al. (2017), who noted barriers to vaccine access primarily among low-education and economically disadvantaged groups. Additionally, a study by Mulu et al. (2021) in Ethiopia found that higher education levels were associated with greater awareness and acceptance of the typhoid vaccine. This variation may be attributed to cultural differences and trust in the healthcare system across the study locations, regardless of participants’ education level. Trust has been referred to as a relational concept, and it is the “backbone” that sustains working relationships in organizations, institutions, and systems, including the health system (Gilson, 2003). An unfavorable or inadequate past relationship experience with or inside a system, particularly including a degree of breach of trust, may lead to disobedience in later initiatives. Instances include; the Tuskegee Syphilis Study, which was ordered to purposefully harm American Blacks (Armstrong, 2008; Williamson & Bigman, 2018), and the allegations of the deliberate injection of children with HIV by volunteer healthcare workers in a hospital in Libya in the 1980s (Rosenthal, 2006). Similarly, the recent violence in the Democratic Republic of the Congo during the Ebola outbreak was misinterpreted as a plot to inject people with lethal drugs because it was heightened by a general mistrust of the democratic system (Maxmen, 2019). In Nigeria, the effect of an improperly implemented health intervention in the northern part of the country resulted in fatalities among study participants and subsequently led to a later rejection of the polio vaccine particularly in that region as a result of a loss of confidence in government to keep them safe (Ghinai, Willott, Dadari, & Larson, 2013; Jegede, 2007). Additionally, a study conducted in Ibadan, Nigeria, investigating the role of trust in enrollment patterns for a social health insurance scheme, found that trust significantly influences the choice of healthcare facilities. The study highlighted that low levels of trust in the government and its policies had a major impact on the acceptance of the National Health Insurance Scheme (NHIS), (Adewole, Reid & Oni, 2021).

## Conclusion

Knowledge of typhoid fever was high among participants, with a majority aware of its symptoms and treatment options. However, awareness of the typhoid vaccine was low, reflecting limited knowledge of preventive measures. Although risk perception was high, misconceptions about transmission and vaccine efficacy persisted, impacting preventive behaviors. Socio-demographic factors, particularly gender and education level, influenced vaccine interest, underscoring the need for targeted educational campaigns. Enhancing public understanding of vaccination and addressing prevalent misconceptions are crucial steps toward improving typhoid prevention efforts in Kano, Nigeria

## Limitations

The study’s limitations include potential sampling bias from a single clinic setting, reliance on self-reported data, and a lack of depth in exploring socio-cultural factors.

## Authors’ contributions

- Nirmal Ravi: The contribution of this author to the manuscript involved the initial conception of research goals and aims, significant oversight, and leadership throughout the planning and execution phases of the research activity and played a critical role in the preparation and refinement of this manuscript.
- Oluwaseunayo Deborah Ayando: The author significantly contributed to the manuscript through the application of rigorous formal techniques, including statistical methodology, to analyze and synthesize the study data effectively.

## Data Availability

All data produced in the present work are contained in the manuscript

## Acknowledgment

We extend our sincere gratitude to the management of EHA Clinics for their invaluable assistance and support, which played a pivotal role in the execution of our study.

